# Exploratory analysis of cumulative fatigue derived from volume and intensity indicators in stage races of professional cycling

**DOI:** 10.1101/2024.11.06.24316801

**Authors:** AS. Machado, C. De la Fuente, A. Javaloyes, M. Moya-Ramón, M. Mateo-March, FP. Carpes

**Affiliations:** Applied Neuromechanics Research Group, Multicenter Graduate Program in Physiological Sciences, Federal University of Pampa, 97500-970 Uruguaiana, RS, Brazil; Exercise and Rehabilitation Sciences Institute, Postgraduate, Faculty of Rehabilitation Sciences, Universidad Andres Bello, Universidad Andres Bello, 7591538 Santiago de Chile, Chile; Sport Sciences Department, Universidad Miguel Hernández de Elche, 03202 Elche, Spain; Sport Sciences Department, University Miguel Hernandez, Alicante Institute for Health and Biomedical Research (ISABIAL-FISABIO Foundation), 03202 Alicante, Spain; Faculty of Sport Sciences, Universidad Europea de Madrid, 28670 Madrid, Spain

**Keywords:** exhaustion, recovery, muscle damage, sports performance, endurance

## Abstract

**Objective:** Here, we discuss cumulative fatigue based on volume and power output from 12 professional male cyclists during two consecutive editions of the Giro d’Italia.

**Methods:** Volume and power output were recorded and described according to time at different intensity zones based on power output (Z0 lower to Z7 higher). Correlations, principal component analysis (PCA), Gaussian clustering, and two-way ANOVA were performed (type error I of 5%).

**Results:** The higher intensity zones elicited higher power output in those shorter stages (R^2^ = 0.54). In contrast, the lower intensity zones were predominant in longer stages. The time spent in Z1 to Z3 (r = 0.67, 0.84, and 0.73) correlated more with stage’s volume duration than time in Z4 to Z7 (r = 0.48, 0.44, and 0.51, and 0.38). The normalized volume declined between stages 2 to 4, 8 to 10, 13 to 15, and 18 to 19. Negative slopes of time spent in Z4 and Z6 occurred one or two stages before Z1 presented negative slopes. In contrast, positive slopes of higher intensity zones were observed with a negative slope of Z1. Different clusters distributions of time volume to complete the grand tour were found (p<0.05). Finally, average power in Z1, Z2, Z3, Z4, Z5, Z6, and Z7 explained 63.62%, 18.20%, 8.12%, 5.84%, 2.98%, 1.20%, and 0.02% of the total variance in the normalized time volume, respectively. Volume and power zone data can recognize cumulative fatigue and performance recovery during a grand tour. Rest days favored the performance recovery, mostly the second rest day.

## Introduction

For professional men cycling athletes, there are three main grand tours with similar characteristics of consecutive days of racing and extreme exigence in terms of performance. The grand tours have a mileage of 3,300 to 3,500 km divided into 21 stages, with single days of rest distributed along the stages. In these races, athletes achieve an average power output of around 260 W [1]. However, little knowledge about stage profiles has been considered [2].

Consecutive days of racing induce cumulative fatigue [1]. Muscle fatigue reduces the ability to produce force over time [3] and causes muscle damage that triggers physiological events i.e. oxidative stress and inflammation [4]. The inflammatory reaction (a cycle lasting up to 120 h) starts restoring the muscle integrity process (muscle homeostasis and regeneration) [5]. Unfortunately, consecutive days of exercise interrupt the recovery cycle and reinforce the production of muscle damage, oxidative stress, and inflammation resulting in the cumulative fatigue condition [6]. Previous studies have explored the effects of cumulative fatigue on isolated muscle conditions [7], in incremental exercise protocols [8], in diverse populations [9], as well as its interactions with other aspects of exercise like psychological and cognitive conditions [10]. However, the description of cumulative fatigue over multistage professional races is difficult to obtain and analyze. Hence the evidence is limited.

In this regard, the current tools of performance monitoring and data sharing can allow the in-field monitoring of athletes’ performance. This is the case of power output reported during the races, which allows a reliable description of the race demands [11]. Therefore, a common way to describe the exercise characteristics during a race is to quantify the volume (time) in different intensity zones according to the power output. These intensity zones can be defined as a percentage of the individual functional power threshold [12,13].

Because the performance in multistage races, like a professional cycling grand tour composed of 21 stages with at least two rest days, generates cumulative fatigue, the distribution of volume between the intensity zones could be altered, denoting a performance change. Finally, these races also include rest days. The rest days can benefit an athlete’s recovery after strong efforts in consecutive days, in which the race is completed spending around 5,000 Kcal/day with high demands for hydration (around 6.7 L/day) [14]. Currently, it is not clear how the variables volume and power output vary in response to rest days or during the race. Therefore, we conducted an exploratory analysis of the time course of cumulative fatigue effects on the volume and power output of professional male cyclists racing during two editions of a grand tour. In addition, we analyzed how the rest days impact cumulative fatigue indicators.

## Material and methods

### Participants and experimental design

Twelve professional cyclists from the same UCI World-Tour professional cycling team and racing two editions of the Giro d’Italia participated in this study. The participants had 26.9 (3.6) years, 178.5 (6.6) cm of height, 69.0 (7.8) kg of body mass, and 21.8 (1.6) kg/m^2^ of body mass index. All signed an informed consent form. This study was approved by the local institution’s ethics committee, and procedures followed the Declaration of Helsinki.

Data collection during the Giro d’Italia 2015 and 2016 included the individual recording of power output and volume distribution for each stage of the race while the athletes followed the planning and protocols defined by the team management. We chose to analyze two editions of the grand tour to make the data generalization more robust. Regarding the characteristics of the race, we considered the number of stages, distance of each stage, and description of each stage considering the information from the official competition website.

### Data collection

Data were recorded using the same device model and configuration for all cyclists (Garmin 510, Garmin Inc., Kansas, United States). The instrument was a power meter, a portable crank-based device (Power2Max type S, Zossen, Germany) that measures the mechanical power considering torque data obtained from an instrumented crankset with strain gages. All power meters were factory calibrated at least once per season, and a zero-offset was performed before each session attending to manufacturers’ instructions. Potential spikes were checked and removed using specific software (Data Spike ID and FIX chart, WKO5 Build 576; TrainingPeaks LLC, Boulder, CO). Hence, volume and power output data were available to be obtained from cyclists’ bicycles. The device measured power output every 1 s with an accuracy of 2% [15]. Finally, when each stage ended, the volume and power output data were uploaded to a cloud service (TrainingPeaks, Boulder, United States) and subsequently, analyses were performed using specific software (WKO5 Build 576; TrainingPeaks LLC, Boulder, CO).

### Intensity zones

Seven exercise intensity zones were determined based on the individual functional threshold power (FTP) [12]: zone 1 (Z1; ≤ 55% of FTP), zone 2 (Z2; between 56 and 75% of FTP), zone 3 (Z3; between 76 and 90% of FTP), zone 4 (Z4; between 91 and 105% of FTP), zone 5 (Z5; between 106 and 120% of FTP), zone 6 (Z6; between 121 and 150% of FTP) and zone 7 (Z7; ≥151% of FTP). Z0 corresponds to the time without pedalling. We determined seven intensity zones representing a wide range of intensities, including aerobic and anaerobic efforts. These intensities distribution is commonly used by these cyclists during training and racing. Zones were determined based on the individualized FTP estimated by the best 20-min power output record [16] obtained during the month before the start of the competition in each of the years. In addition, whether cyclists recorded higher 20-min power during the competition, the intensity zones were updated to accurately quantify the intensities distribution.

### Data analysis

We described the time-normalized stages’ duration, which made all stage times equal to 100% in the time domain. Data (time volume normalized by stages duration and power output) were described for each intensity zone as mean and standard deviation considering the normal data distribution verified with Shapiro-Wilk and Levene’s tests with error type 1 equal to 5%. The associations between the time spent in the different intensity zones and the stage duration were described using a non-linear fitting with an exponential series [f_(x)_= a*e^(bx)^ + c*e^(dx)^ + error]. The coefficients, 95%CI (confident interval), and the determination coefficient (R^2^) were described. The association between total time volume and each stage duration was described using a linear model [f_(x)_ = mx + f + error] to identify whether higher or lower slopes of intensity zones are dependent on the period stages (positive slopes >0.1 vs constants <0.1). The slope (m), inclination angle (θ), correlation and determination coefficient (R^2^) were described. The peak of higher declines in the normalized volume was measured as the negative slopes across stages before the next normalized volume showed an increment (positive slopes). The slope changes in normalized time were tracked in the zone intensity with the highest dispersion that could show the compromise of the time-volume output. Additionally, the intensity increases and decreases strategies along a stage were explored by the slope changes normalized by their magnitudes (absolute values), resulting in −1 (negative slope) or +1 (positive slope) indicators. Finally, the principal component analysis (PCA) was used to understand how the intensity zones (Z1, Z2, Z3, Z4, Z5, Z6, and Z7) explained the total variance of normalized time volume for each stage. A threshold of 80% of the total variance was used to choose the principal component and understand the structure of the data. In addition, a mixed Gaussian fitting was used to define data clusters in the principal component space. The Akaike information criterion was used to determine the number of components [AIC = 2*k* – 2ln(*L*)], with *k* equal to the number of clusters and *L* equal to the maximum value of the likelihood function for the Gaussian model. The posterior probability of pertinence of the main cluster fitted by mixed Gaussian distributions was described, and the normalized time volume and power output for each found cluster was also described. Finally, the clusters for normalized time and power output were described and compared using a two-way ANOVA and multiple comparisons with error type I of 5%.

## Results

Table 1 describes the main characteristics of the Giro d’Italia stages for each year analyzed.

**Table 1.** Descriptive information from each analyzed Giro d’Italia edition.

| 2015 |  | 2016 |  |
| --- | --- | --- | --- |
| Stage: location<br>Date, distance<br>Won how | Vertical meters<br>Average speed winner<br>Average power<br>Average cadence | Stage: location<br>Date, distance<br>Won how | Vertical meters<br>Average speed winner<br>Average power<br>Average cadence |
| 1: San Lorenzo a Mare<br>Saturday, May 9, 17.6 km<br>Time trial | 83 m<br>54.34 km/h<br>355.86 W<br>94.69 rpm | 1: Apeldoorn ITT<br>Friday, May 6, 9.8 km<br>Time trial | 37 m<br>53.21 km/h<br>378.6 W<br>93.24 rpm |
| 2: Albenga → Genova<br>Sunday, May 10, 177 km<br>Sprint of large group | 1868 m<br>41.93 km/h<br>190.21 W<br>73.08 rpm | 2: Arnhem → Nijmegen<br>Saturday, May 7, 190 km<br>Sprint of a large group | 732 m<br>40.93 km/h<br>153.9 W<br>62.67 rpm |
| 3: Rapallo → Sestri Levante<br>Monday, May 11, 136 km<br>Sprint of large group | 2797 m<br>38.15 km/h<br>222.70 W<br>74.42 rpm | 3: Nijmegen → Arnhem<br>Sunday, May 8, 190 km<br>Sprint of a large group | 691 m<br>43.22 km/h<br>184.2 W<br>70.46 rpm |
| 4: Chiavari → La Spezia<br>Tuesday, May 12, 150 km<br>14.1 km solo | 3050 m<br>39.48 km/h<br>231.68 W<br>74.59 rpm | 4: Catanzaro → Praia a Mare<br>Tuesday, May 10, 200 km<br>9 km solo | 2418 m<br>41.83 km/h<br>217.6 W<br>72.77 rpm |
| 5: La Spezia → Abetone<br>Wednesday, May 13, 152 km<br>10.7 km solo | 3066 m<br>36.58 km/h<br>216.86 W<br>71.95 rpm | 5: Praia a Mare → Benevento<br>Wednesday, May 11, 233 km<br>Sprint of a large group | 3459 m<br>41.05 km/h<br>202.9 W<br>66.54 rpm |
| 6: Montecatini Terme →<br>Castiglione della Pescaia<br>Thursday, May 14, 183 km<br>Sprint of large group | 1625 m<br>42.28 km/h<br>205.16 W<br>72.51 rpm | 6: Ponte → Roccaraso<br>Thursday, May 12, 157 km<br>15 km solo | 3749 m<br>33.63 km/h<br>226.2 W<br>70.97 rpm |
| 7: Grosseto → Fiuggi<br>Friday, May 15, 264 km<br>Sprint of a large group | 2873 m<br>35.81** km/h<br>193.73 W<br>68.79 rpm | 7: Sulmona → Foligno<br>Friday, May 13, 211 km<br>Sprint of a large group | 2301 m<br>42.04 km/h<br>223.2 W<br>69.85 rpm |
| 8: Fiuggi → Campitello Matese<br>Saturday, May 16, 186 km<br>3.4 km solo | 3842 m<br>38.28* km/h<br>241.91 W<br>75.02 rpm | 8: Foligno → Arezzo<br>Saturday, May 14, 186 km<br>24.6 solo | 1985<br>43.92<br>255.8<br>79.46 |
| 9: Benevento → San Giorgio del Sannio<br>Sunday, May 17, 215 km<br>4 km solo | 4264 m<br>38.34 km/h<br>259.87 W<br>74.34 rpm | 9: Chianti ITT<br>Sunday, May 15, 40.5 km<br>Time trial | 561 m<br>46.96<br>353.6<br>86.23 |
| 10: Civitanova Marche → Forlì<br>Tuesday, May 19, 200 km<br>Sprint of a small group | 841 m<br>45.07 km/h<br>206.70 W<br>72.81 rpm | 10: Campi Bisenzio → Sestola<br>Tuesday, May 17, 219 km<br>14.5 km solo | 4618<br>38.14<br>265.2<br>72.31 |
| 11: Forlì → Imola<br>Wednesday, May 20, 153 km<br>25 km solo | 2949 m<br>39.04 km/h<br>206.70 W<br>72.81 rpm | 11: Modena → Asolo<br>Wednesday, May 18, 229 km<br>Sprint of a small group | 777 m<br>45.93<br>215.8<br>72.12 |
| 12: Imola → Vicenza<br>Thursday, May 21, 190 km<br>Sprint of a large group | 1288 m<br>43.37 km/h<br>212.21 W<br>74.98 rpm | 12: Noale → Bibione<br>Thursday, May 19, 182 km<br>Sprint of a large group | 122 m<br>42.66<br>196.2<br>71.82 |
| 13: Montecchio Maggiore → Jesolo<br>Friday, May 22, 147 km<br>Sprint of a large group | 131 m<br>48.16 km/h<br>239.48 W<br>74.80 rpm | 13: Palmanova → Cividale del Friuli<br>Friday, May 20, 170 km<br>33.4 km solo | 3669<br>37.53<br>272.3<br>71.89 |
| 14: Treviso → Valdobbiadene (ITT)<br>Saturday, May 23, 59.2 km<br>Time trial | 593 m<br>45.77 km/h<br>295.88 W<br>82.78 rpm | 14: Alpago → Corvara<br>Saturday, May 21, 210 km<br>Sprint of a small group | 6001<br>34.4<br>253.4<br>70.54 |
| 15: Marostica → Madonna di Campiglio<br>Sunday, May 24, 165 km<br>Sprint of a small group | 4634 m<br>37.70 km/h<br>325.41 W<br>81.13 rpm | 15: Castelrotto → Alpe di Siusi<br>ITT<br>Sunday, May 22, 10.8 km<br>Time trial | 785 m<br>22.06<br>389.3<br>81.23 |
| 16: Pinzolo → Aprica<br>Tuesday, May 26, 174 km<br>4 km solo | 4951 m<br>35.07 km/h<br>257.25 W<br>73.50 rpm | 16: Bressanone → Andalo<br>Tuesday, May 24, 132 km<br>Sprint a deux | 2737<br>44.27<br>289.9<br>75.64 |
| 17: Tirano → Lugano (CH)<br>Wednesday, May 27, 134 km<br>Sprint of a small group | 1857 m<br>42.80 km/h<br>211.83 W<br>73.26 rpm | 17: Molveno → Cassano d'Adda<br>Wednesday, May 25, 196 km<br>0.4 km solo | 1567<br>43.32<br>206.3<br>67.17 |
| 18: Melide (CH) → Verbania<br>Thursday, May 28, 170 km<br>19.3 km solo | 2195 m<br>41.76 km/h<br>212.96 W<br>74.82 rpm | 18: Muggio → Pinerolo<br>Thursday, May 26, 244 km<br>Sprint of a small group | 1637<br>44.97<br>211.5<br>71.84 |
| 19: Gravellona Toce → Cervinia<br>Friday, May 29, 236 km<br>6 km solo | 5113 m<br>36.85 km/h<br>252.22 W<br>74.32 rpm | 19: Pinerolo → Risoul<br>Friday, May 27, 162 km<br>5.1 km solo | 3905<br>37.40<br>284.7<br>74.46 |
| 20: Saint Vincent → Sestriere<br>Saturday, May 30, 196 km<br>1.6 km solo | 3355 m<br>37.64 km/h<br>230.75 W<br>72.45 rpm | 20: Guillestre → Sant'Anna di<br>Vinadio<br>Saturday, May 28, 134 km<br>14.1 km solo | 4500<br>30.60<br>262.9<br>62.45 |
| 21: Torino → Milano<br>Sunday, May 31, 185 km<br>Sprint a deux | 142 m<br>42.92 km/h<br>172.54 W<br>72.50 rpm | 21: Cuneo → Torino<br>Sunday, May 29, 163 km<br>Sprint of a large group | 879 m<br>42.84 |
| Team athletes in the top 10<br>classification | 1 |  | 2 |

\* Winner was an athlete of the team. \*\* second place was an athlete of the team. \*\*\* third place was an athlete of the team. Source: PRO Cycling stats (https://www.procyclingstats.com/race/giro-d-italia/2015/stage-1/result/result; https://www.procyclingstats.com/race/giro-d-italia/2016/stage-1/result/result).

Cyclists spent more time (% stage time) in Z1, followed by Z2, Z3, Z0, Z4, Z5, Z6, and Z7. The average time spent (s), mean (standard deviation) in Z0, Z1, Z2, Z3, Z4, Z5, Z6, and Z7 were 14.6 (5.8), 26.3 (12.6), 17.3 (6.2), 14.7 (6.4), 13.0 (10.2), 7.3 (6.3), 4.8 (3.0), and 2.1 (1.6) % stage duration, respectively. The most variable zone was Z1, followed by Z4, Z3, Z5, Z2, Z0, Z6, and Z7. The mean (standard deviation) power output (expressed in Watts) in each of the intensity zones for Z0, Z1, Z2, Z3, Z4, Z5, Z6, and Z7 was 122.9 (13.9), 247.5 (17.2), 311.1(21.4), 366.0 (24.9), 421.1 (28.5), 496.5 (33.8), and 658.2 (47.6) Watts, respectively.

The association between mean power and stage time was non-linear modeled [y = 334.8 [95%CI 238.7-430.9] e^(−0.0007997^ ^[95%CI-0.001061^ ^+^ ^0.0005382]^ ^x)^ + 306.8 [95%CI 204.2 - 409.5] e^(−0.0001052^ ^[95%CI-0.000163^ ^+^ ^0.00004739]^ ^x)^ R^2^ = 0.54]. The highest intensity zones are related to longer time sustained at higher power output levels in shorter stages. In contrast, as Figure 1A depicts, the lower intensity zones were used to cover stages with longer durations, developing lower average power during the two analyzed editions of the Giro d’ Italia.

**Figure 1.**
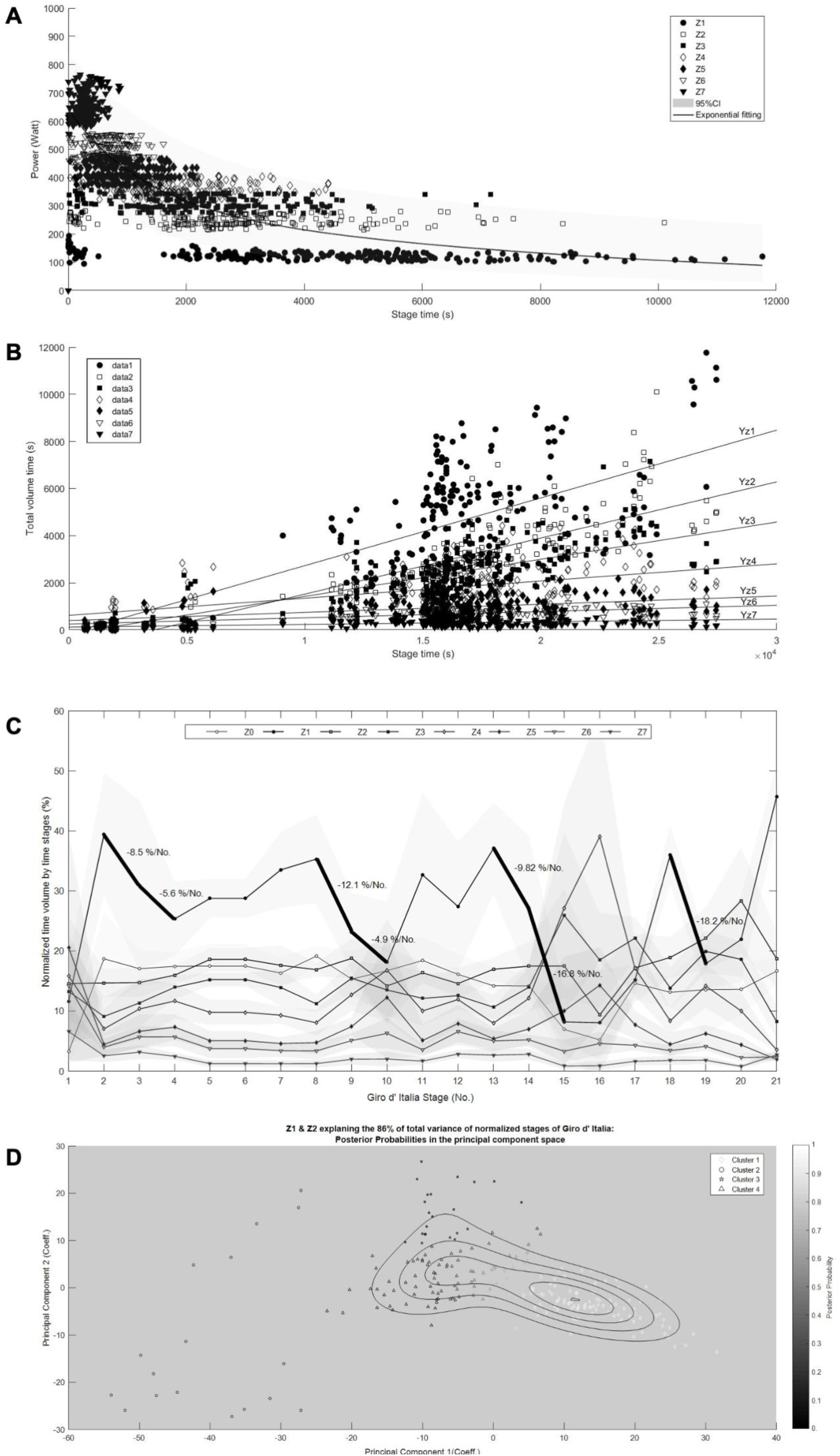
**A:** Power vs stage time for intensity zones entitled Z1 to Z7 during two editions of the Giro d’ Italia. The power was measured in Watts and the stage time was measured in seconds. The figure shows the non-linear association between power and time during two editions of the Giro d’ Italia. The highest intensity zones were predominant in stages of shorter duration requesting athletes to develop higher average power, while the lower intensity zones were predominant in longer stages requiring lower average power. Note the double distribution of Z1 and Z2 that suggest the lowest race period predominantly involve time in Z1 and Z2 intensity zones. **B:** Total volume time vs stage time for intensity zone Z1 to Z7 during two editions of the Giro d’ Italia. The intensity zones are entitled Z1, Z2, Z3, Z4, Z5, Z6, and Z7, and each slope is entitled Yz1, Yz2, Yz3, Yz4, Yz5, Yz6, and Yz7, respectively. The total volume time was measured in seconds, and each stage duration was measured in seconds. **C:** Normalized time volume by time stage during the Giro d’ Italia. The intensity zones are entitled Z0, Z1, Z2, Z3, Z4, Z5, Z6, and Z7. The mean and standard deviation are shown with black lines and gray shadows, respectively. The thick lines show the higher decrease of time volume tracked in Z1, the intensity zone with the most dispersion. The rest days occurred between stages 9 and 10, and stages 14 and 15. The first rest day of 2016 edition was not considered. **D:** The posterior probability of cluster pertinence of the main Gaussian cluster (cluster 1) in the principal component space explained 86% of the total data variance. Four clusters were identified in the principal component space from the Akaike information criterion [AIC = 2*k* – 2ln(*L*) with *k* equal to the number of clusters and *L* equal to the maximum value of the likelihood function for the Gaussian model].

The association between total time volume (time to finish the grand tour) and the time volume for each stage (Figure 1B) showed higher slopes and inclination angles for Z1 [m = 0.29, θ = 16.2°, R^2^ = 0.45], followed by Z2 [m = 0.24, θ =13.5°, R^2^ = 0.71], Z3 [m = 0.16, θ =9.1°, R^2^ = 0.53], Z4 [m = 0.08, θ = 4.6°, R^2^ = 0.23], Z5 [m = 0.04, θ =2.3°, R^2^ = 0.20], Z6 [m = 0.03, θ =1.7°, R^2^ = 0.26], and Z7 [m = 0.01, θ = 0.6°, R^2^ = 0.15].

The analysis of mean normalized time spent in the different intensity zones across each of the 21 stages from each year edition (Figure 1C) showed that higher peak declines of normalized volume, measured as the negative slopes, across stages before the next increment that occurred between stages 2 and 4 [Δ= −8.5 % No.^-1^], 8 and 10 [Δ= −12.2 % No.^-1^], 13 and 15 [Δ= −19.0 % No.^-1^], and 18 and 19 [Δ= −18.2 % No.^-1^] tracked in the zone intensity with the highest dispersion (Z1).

Higher peak slope increment of normalized volume by stage until the next slope decrease after stage 4 [Δ= 4.8 % No.^-1^, between 4 and 8], after stage 10 [Δ= −4.6 % No.^-1^, between 10 and 13], after stage 15 [Δ= 20.8 % No.^-1^, between 15 and 18], and after stage 19 [Δ= 23.8 % No.^-1^, between 19 and 21] tracked in the zone intensity with the highest dispersion (Z1), see Figure 1C.

Negative slopes of Z3, Z4, Z5, and Z6 occurred four times and three times in Z7 before one or two stages before Z1 presented negative slopes (Table 2). In contrast, positive slopes of higher intensity zones (Z3 three times, Z4 four times, Z5 four times, Z6 three times, and Z7 twice during the competition) were observed, accompanied by a negative slope of Z1 (Table 2).

**Table 2.** Slope changes of normalized mean volume patterns across the *Giro d’ Italia*.

|  | Giro d' Italia Stages |  |  |  |  |  |  |  |  |  |  |  |  |  |  |  |  |  |  |  |
| --- | --- | --- | --- | --- | --- | --- | --- | --- | --- | --- | --- | --- | --- | --- | --- | --- | --- | --- | --- | --- |
|  | 1 | 2 | 3 | 4 | 5 | 6 | 7 | 8 | 9 | 10 | 11 | 12 | 13 | 14 | 15 | 16 | 17 | 18 | 19 | 20 |
| Z0 | 1 | -1 | 1 | 1 | 0 | -1 | 1 | -1 | 1 | 1 | -1 | -1 | -1 | -1 | -1 | 1 | -1 | 1 | 1 | 1 |
| Z1 | 1 | -1 | -1<sup>R</sup> | 1 | 0 | 1 | 1 | -1 | -1<sup>R</sup> | 1 | -1 | 1 | -1 | -1 | -1<sup>R</sup> | 1 | 1 | -1 | 1 | 1 |
| Z2 | 1 | 1 | 1 | 1 | 0 | -1 | -1 | 1 | -1 | 1 | -1 | 1 | 1 | 1 | -1 | 1 | 1 | 1 | 1 | -1 |
| Z3 | -1 | 1 | 1 | 1 | 0 | -1 | -1 | 1 | -1 | -1 | 1 | -1 | 1 | 1 | -1 | 1 | -1 | 1 | -1 | -1 |
| Z4 | -1 | 1 | 1 | -1 | 0 | -1 | -1 | 1 | 1 | -1 | 1 | -1 | 1 | 1 | 1 | -1 | -1 | 1 | -1 | -1 |
| Z5 | -1 | 1 | 1 | -1 | 0 | -1 | 1 | 1 | 1 | -1 | 1 | -1 | 1 | 1 | 1 | -1 | -1 | 1 | -1 | -1 |
| Z6 | -1 | 1 | 1 | -1 | 0 | -1 | -1 | 1 | 1 | -1 | 1 | -1 | 1 | -1 | 1 | -1 | -1 | 1 | -1 | 1 |
| Z7 | -1 | 1 | -1 | -1 | 0 | -1 | 1 | 1 | 1 | -1 | 1 | -1 | 1 | -1 | 1 | 1 | 1 | 1 | -1 | 1 |

R = rest stage
-1 = negative slope (volume increase)
1 = positive slope (volume decrease)
Z0 = intensity zone zero
Z1 = intensity zone one
Z2 = intensity zone two
Z3 = intensity zone three
Z4 = intensity zone four
Z5 = intensity zone five

The principal component analysis showed that time spent in intensity zones Z1, Z2, Z3, Z4, Z5, Z6, and Z7 explained 63.62%, 18.20%, 8.12%, 5.84%, 2.98%, 1.20%, and 0.02% of the total variance in the normalized time by each time stage, respectively. The best number of mixed Gaussian components was four from the Akaike information criterion. The posterior probability of pertinence of the main cluster fitted by mixed Gaussian distributions in the principal component space that explained 86% of the total variance is summarized in Figure 1D.

There was a main effect for clusters and intensity zones in normalized time-volume (p<0.001), and there was interaction (p<0.001). Clusters one and two differed (p<0.001), as well as clusters one and four differed (p<0.001). There were differences between all intensities (p<0.001, Table 3). There was a main effect for intensity zones in power (p<0.001) without interaction (p>0.05) or effect for clusters (p>0.05). All intensity zones differed (p<0.001, Table 3).

**Table 3.**
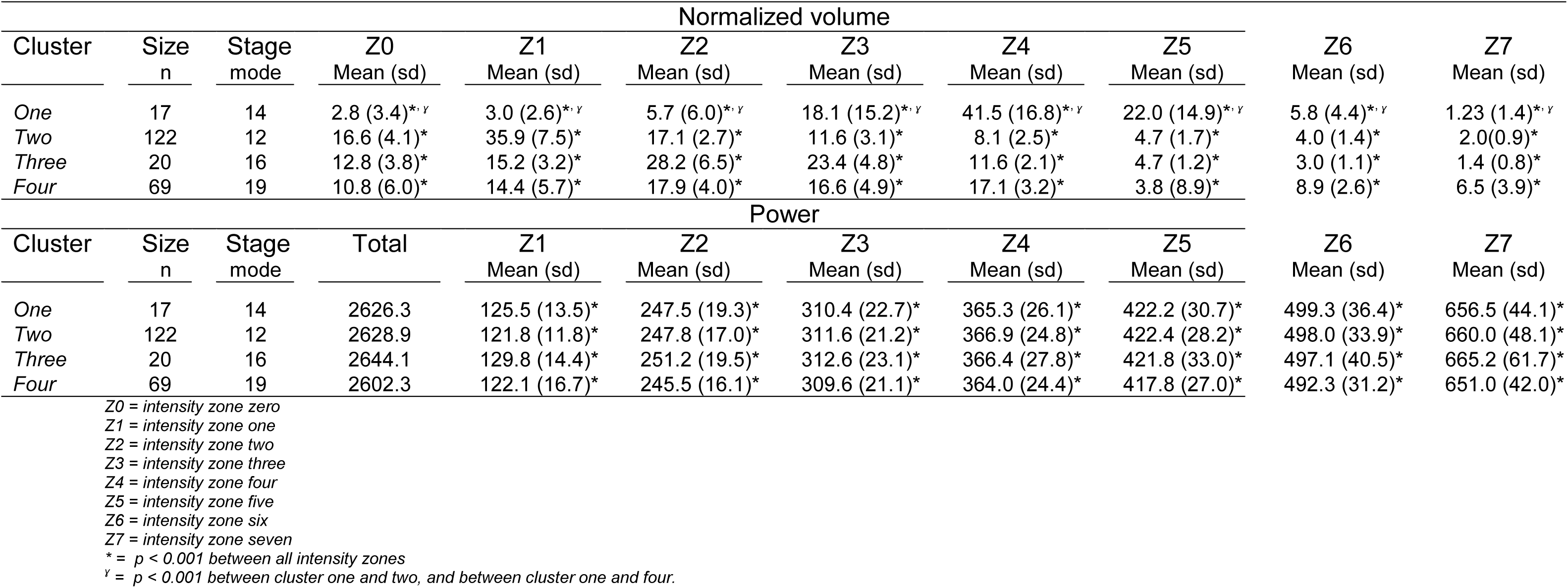
Volume and power cluster differences.

## Discussion

A strong novelty of our study is that the analysis of two grand tour editions allowed the verification of cumulative fatigue during consecutive days of racing at a high level of competition. Importantly, cumulative fatigue has negative effects on performance and can cause overuse muscle injuries (26), the third largest cause of professional athlete absence in Grand Tours [17]. Our main findings here were that cyclists i) develop higher mean power during shorter time stages using higher intensity zones and develop lower mean power during longer time stages using lower intensity zones, ii) use lower intensity zones to cover long-time stages while higher intensity zones are used to cover lower-time stages, iii) experienced cumulated fatigue during the competition (there was four recognizable performance decreases indicating fatigue effects) and the second rest day had the best impact on performance increase for subsequent stages, iv) decrease the time-volume of Z3, Z4, Z5, Z6 and Z7 in two or three stages before the performance decrease, which suggests those intensities being predictors, while higher zone intensities suggest compensating the performance decrease, and v) develop almost two different intensity zones distribution (clusters) in the time-volume to complete the competition without power generation differences.

Our findings have applicability in sports because high mileage cycling events such as Giro d’Italia, Vuelta a España, and Tour de France have similar energy expenditure and intensity development [18]. We consider our findings can help both specific race strategies during the Giro d’Italia and planning training. The grand tours involve several consecutive days of high-level physical effort [19]. Thus, adequate training planning should consider not only experimental laboratory knowledge but also in-field race characteristics and performance analysis because not always the in-field conditions are possible to reproduce fully in laboratory testing [20].

Along the two editions of the Giro, we found how the stage distances determine the distribution of effort zones. Accordingly, longer stages use mainly lower intensities that represent low-intensity exercise below the first lactate or ventilatory threshold [21]. Lower intensity zones showed how relevant they were to be completing the stages. Even shorter stages have shown activity in lower zone intensities (double distribution for Z1 to Z3) in coherence with the maximum efforts cannot be sustained for the entire stage because fatigue will limit performance (high-intensity exercise over the lactate or ventilatory threshold [22]), either by peripheral or central pathways [23,24]. Thus, higher intensities are chosen for shorter time stages during the Giro d’ Italia in accordance that each stage’s intensity is modulated by total race duration in elite cycling [25].

Therefore, it is crucial to make an appropriate choice of the mechanical power provided by cyclists (mechanical power being defined by: the aerodynamic friction, the ascending term, and the accelerating term) in each stage of the competition to increase the cyclist’s performance [26]. Hence, the adequate use of intensity zones during the competition directly impacts the cycling dynamics. This choice suggests helping prevent fatigue episodes and increased fatigue intensity (lower cumulated fatigue) when the mechanical power is opportunely delivered. Likely, intensified training strategies may favor power production that will improve critical moments of the tour [27] complemented with aerobic exercise intensity [28], and might explain why shorter stages seem to be decisive for cyclist performance in the race [1].

Regarding cumulative fatigue, there were four recognizable performance decreases (negative slopes during the time-volume across the stages). The negative slopes tracked in the most variable intensity zone, which explained the main variance of the total data studied by principal component analysis [29], could be interpreted as the incapacity to sustain a stable peripheral effort in low-intensity zones. The main cause of performance losses in repetitive tasks is caused by cumulative fatigue [6]. The high correlation between blood lactate transition thresholds and endurance performance, and the aerobic and anaerobic indices of cycling performance [30] support our effort to track the decrease in cycling performance in the time course of the competition. However, It is also possible that performance decreases allow athletes to save energy, especially for the final moments of a race, when habitually the classification tends to be decided [31].

In our exploration, three of four slope decreases fitted with the given rest stages (stage 3 in 2016, stage 9 in 2015 and 2016, and stage 15 in 2015 and 2016), showing an immediate reversion after the rest day. This influence of the rest day suggests the ability to choose to stay longer at higher intensity zones (except for the opening stage). Therefore, we argue that the variation in the distribution of intensities over time can describe the process of accumulated fatigue installation and the effect of rest days on time at different intensity zones. In addition, we also observed a decrease in time-volume in Z3, Z4, Z5, Z6, and Z7 in two or three stages before the drop in performance. These factors need to be further studied to be discussed as possible performance predictors. In the same way, time-volume at higher zone intensities increased when the performance drops, which seems to be a compensation strategy during the competition. Finally, there were almost two intensity zone pattern distributions in the time volume to complete the competition without differences in the power delivery. The main percentual differences were in Z1, Z4, and Z5, where one cluster completed the competition with a lower time in Z1 but stayed more time in Z4 and Z5. Zone intensities 4 and 5 permitted production of more than 320 W and lower than 450 W. It suggests a strategy that favors the use of intensity zones that can deliver satisfactory values of power. This kind of pattern may risk more cumulative fatigue if the athletes do not have enough preparation or induce more muscle damage, oxidative stress, and inflammation events during the competition. Therefore, the surveillance of how the time-volume in different intensity zones is used, the athlete’s characteristics, the race conditions, and the athlete’s interaction with these conditions are crucial to define adequate planning training and evaluate if the athletes are under risk cumulative fatigue conditions during the race, decreasing the athlete’s and team performance.

Our study has limitations. The first one is the fact that our participants are all men, and we know that the behavior of women for these same tests is different [32]. Therefore, the extrapolation of these results to female athletes is not possible. In addition, we know that different athletes perform specific tasks within the strategy of a team in the competition and that this specialization generates differences in performance in the tests for each athlete [33]. Finally, we also recognize that a real-world condition for data collection is subject to confounding factors that rely on the variables mentioned, but also the motivation of the athletes, soreness, effort perception, and individual goals. Despite these limitations, we consider that the uniqueness of our exploratory analysis ensures sufficient novelty and relevance to our study.

## Data Availability

All data produced in the present study are available upon reasonable request to the authors

